# Impact of bystander and dispatcher interventions on clinical outcomes in Out-of-Hospital Cardiac Arrest: A Study of 8,253 Polish Cases from the European Registry of Cardiac Arrest

**DOI:** 10.1101/2025.01.05.24319824

**Authors:** Anna Żądło, Dawid Wilczek, Grzegorz Cebula, Daria Orzechowska, Tomasz Tokarek

## Abstract

**INTRODUCTION:** Out-of-hospital cardiac arrest (OHCA) remains a significant public health challenge. This study aimed to assess impact of bystander and dispatcher interventions on survival outcomes in OHCA cases in Poland using data from the European Registry of Cardiac Arrest (EuReCa).

**MATERIAL AND METHODS:** A retrospective analysis was conducted on 8,253 patients included in the EuReCa between 2014 and 2022. The data was collected from emergency medical team reports, and survival outcomes at 30 days were obtained from the national death registry.

**RESULTS:** A gradual increase in CPR attempted by EMS or bystander, as well as CPR performed with the support of a dispatcher, was observed throughout consecutive years. Dispatcher-guided resuscitation was linked with increased return of spontaneous circulation (ROSC) (22.1% vs 30.6%; *P* =0.01). Patients treated with implementation of telephone-assisted cardiopulmonary resuscitation were associated with a higher prevalence of ROSC (21.0% vs. 27.4%; *P* =0.01).

**CONCLUSIONS:** Dispatcher and bystander interventions significantly improve immediate outcomes in OHCA. However, long-term survival seems to not be improved. System-level strategies to increase AED accessibility, implement real-time support, and standardize Utstein-based reporting in Poland are crucial to increase survival and enable international comparisons.

## INTRODUCTION

Out-of-Hospital Cardiac Arrests (OHCA) present a significant challenge for healthcare systems. The European Resuscitation Council (ERC) initiated the European Registry of Cardiac Arrest (EuReCa) study, involving multiple European countries to collect and analyze data [1]. Notably, it has been proven that with each minute of delay in defibrillation, the probability of survival decreases by 10% to 12% [2]. Previous studies on the epidemiology of OHCA in Poland indicate that bystanders often fail to correctly recognize cardiac arrests, and many are unable to initiate cardiopulmonary resuscitation (CPR). Only a small percentage receive telephone-assisted CPR instructions from dispatchers [3,4]. Some studies have examined bystander interventions and AED utilization, however they are limited to specific regions of the country [3,5]. Additionally, many existing studies focus on the impact of CPR and other emergency medical services interventions while neglecting the role of bystanders [6]. Despite numerous initiatives aimed at improving the Polish EMS system - such as the two-time revision of the ambulance medical emergency procedure report used in Poland (Karta Medycznych czynnosci ratunkowych - KMCR), implementation of new medical dispatcher protocols and increased AED acquisition - gaps remain that weaken the system, and technological support for bystanders remains inadequate. The AED registry lacks synchronization, and reporting on AED usage is minimal [7,8]. Crucially, there is no data on neurological and post-discharge outcomes, with available information only detailing the timing of patient deaths. There is a lack of data from national authorities providing comprehensive analysis on this topic. Standardized outcome reporting is essential to assess long-term survival and guide improvements in post-arrest care. The study by Orzechowski et al. shows that a robust methodology - including survival analysis and structured patient follow-up - can be effectively implemented in Poland to assess the impact of therapeutic home-based interventions [9]. Thus, we sought to investigate the impact of bystander and dispatcher interventions on OHCA outcomes in Poland, addressing existing data limitations and providing insight into potential improvements in the chain of survival.

## MATERIAL AND METHODS

The comprehensive retrospective analysis was conducted on all patients (n=8253) included in the EuReCa study in Poland across three editions between 2014-2022. EuReCa One (one month - October 2014) used paper-based, voluntary data collection prepared by Polish Resuscitation Council representatives and the anonymized National Death Registry.

The whole study was registered on ClinicalTrials.gov (T02236819). EuReCa Two (1st October to 31st December 2017) continued with the same methods and data sources, and was also registered on ClinicalTrials.gov (NCT03130088). EuReCa Three (1st September to 30th November 2022) introduced electronic, mandatory data collection, including patient ambulance records, and is registered with the German Registry of Clinical Trials (DRKS00028591), searchable via the World Health Organization meta-registry [10].

Data was collected from the documentation completed by emergency medical teams at the scene of an OHCA. Due to significant discrepancies in the data across all three editions, the definitions from EuReCa One were adopted to ensure a consistent evaluation of the collected information. All categories introduced in the second and third editions were excluded. Among the missing information are the time of patient discharge from the hospital, neurological outcome, the age of witnesses, the individual who initiated CPR, and details regarding deaths confirmed at the scene.

Considering these factors, further analysis of the data was rendered impossible, limiting the results obtained. Information about 30-day survival was anonymized and obtained by the Air Ambulance Service from the national death registry.

The data collection framework imposed by the authors of the EuReCa study limited access to many variables, while also allowing the use of entries labeled as “unknown” and “not recorded”. Researchers from all participating countries encountered similar challenges, which constitutes one of the study’s limitations. We excluded all data labeled as “unknown” or “not recorded” and focused on analyzing factors related to the role of bystanders in OHCA.

There is currently no national systematic registry for OHCA data in Poland. The EuReCa One and EuReCa Two data collection was facilitated through a registry created by Prof. Grzegorz Cebula connected with the Polish Resuscitation Council specifically for these research purposes. The EuReCa Three data reporting was challenging and required manual analysis based on descriptive records, in order to be able to gather as much information as possible. Specifically, the study examines the actions of bystanders, including emergency recognition, activation of EMS, reception of telephone-assisted CPR (T-CPR) instructions, identification of shockable rhythm and performance of CPR.

The study was provided in congruence with ethical principles derived from the Declaration of Helsinki with later amendments. No financial support was provided for this registry. The protocol was approved by the local ethics committee of the Jagiellonian University, and registered under the number 118.0043.1.299.2024.

The statistical analysis was done with the MedCalc v.18 statistical software (MedCalc Software, Ostend, Belgium). Standard descriptive statistics were used in the analysis. The normality of the data was assessed with the Shapiro-Wilk test. We expressed the categorical variables as numbers and percentages. Quantitative variables were described using their median, supplemented by interquartile ranges. The level of statistical significance was set at *P*<0.05. A direct comparison between the groups was done using the Chi-square test for categorical variables. The one-way ANOVA with post hoc Dunn’s test was used for normal distributions with equal variance between groups, while the Kruskal–Wallis test was used for non-normally distributed data.

## RESULTS

Population characteristics and differences between EuReCa editions are shown in Table I A significant increase in CPR performed with the support of the dispatcher was seen across all editions of the EuReCa Studies (Table I). Likewise, a systematic increase in frequency of shockable rhythm was demonstrated (Table I). The highest incidence of return of spontaneous circulation till hospital arrival (ROSC) was noticed in EuReCa One study, where the proportion of successful CPRs amounted to 39.7%. The worst results were observed in the second edition of the study (Table II). A significant decrease in 30-day survival, from 37.5% and 32.7% in the first and second editions, respectively, to 6.9% in EuReCa Three, was observed (*P*=0.01). Since the first EuReCa study in 2014 a significant increase in the rate the performed CPR was observed (*P*=0.01), especially in the third edition where 95.7% of the OHCA were attempted by bystander or EMS (Table I).

**Figure 1:**
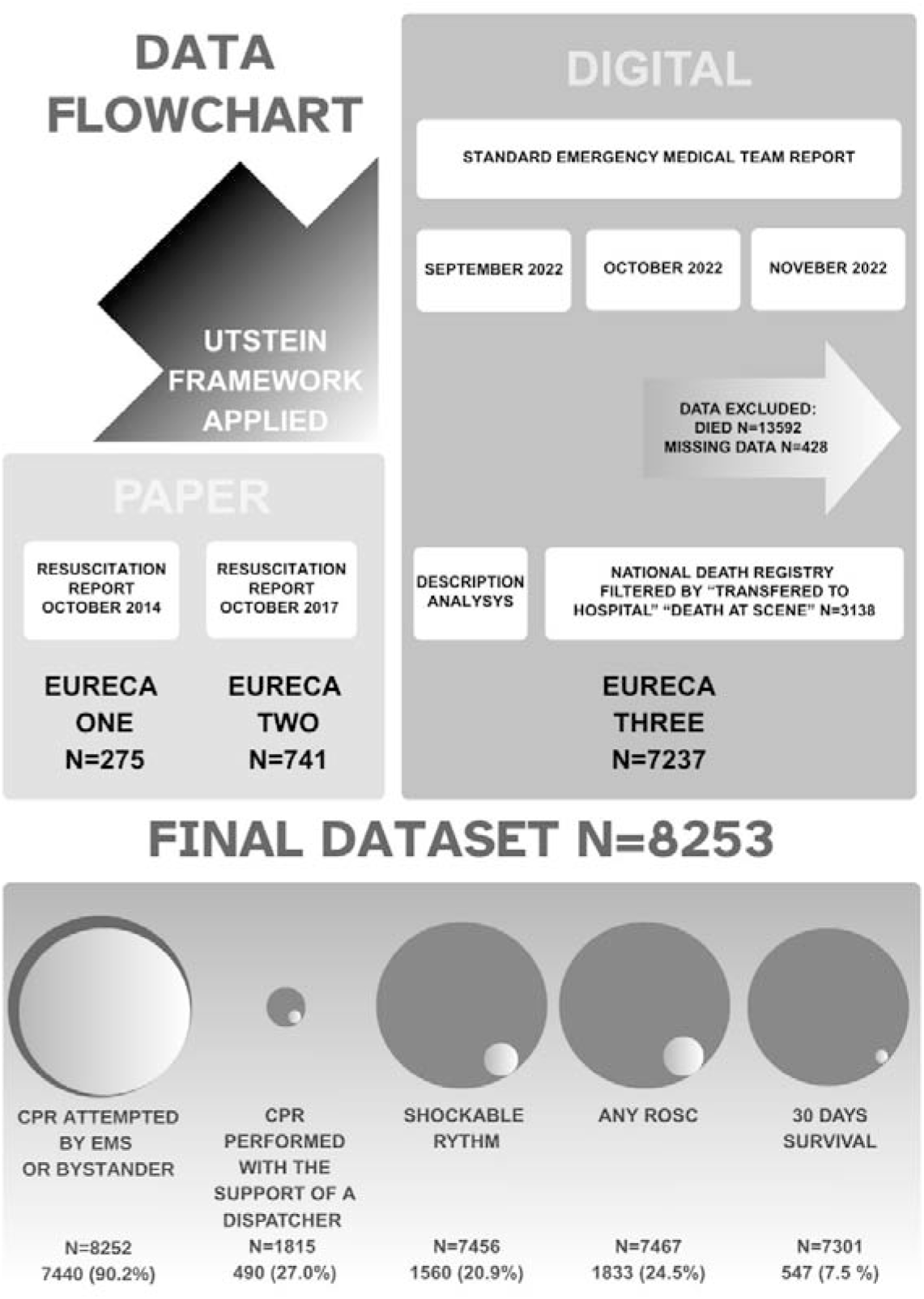
Data flowchart. CPR, cardiopulmonary resuscitation; EMS, emergency medical services; EuReCa, European Registry of Cardiac Arrest; Any ROSC, return of spontaneous circulation till hospital arrive.

**Figure 2:**
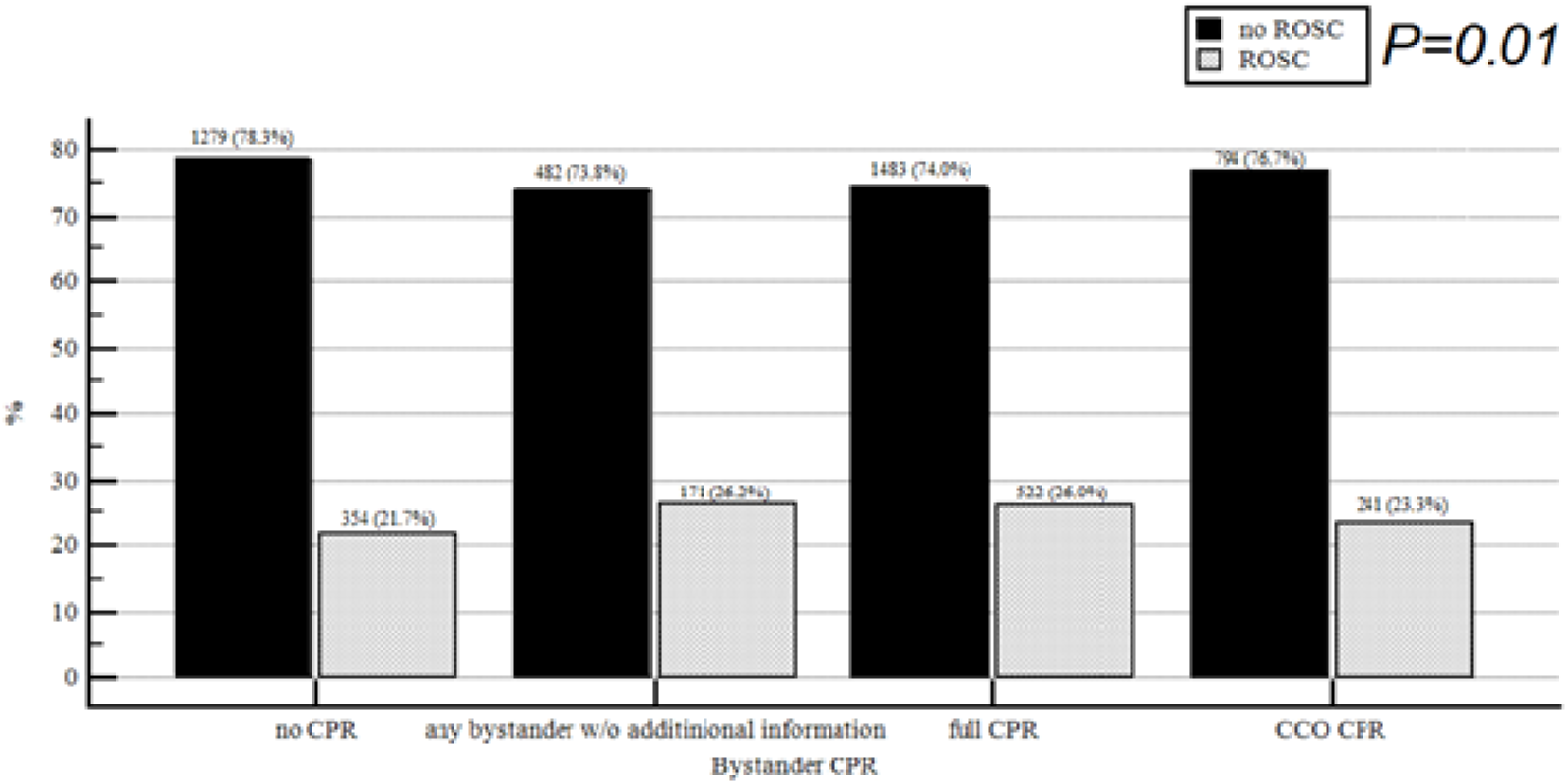
Contribution of Bystander-Assisted Cardiopulmonary Resuscitation in EuReCa Studies. NO CPR, instances where a bystander did not initiate any resuscitation efforts; Any bystander w/o any information, resuscitation provided by a bystander, but no specific details about the type, quality, or extent of the intervention are available; Full CPR, cases where complete resuscitation techniques, including chest compressions and ventilations, were performed; CCO CPR, cases where only chest compressions were performed without ventilation.

**Table I:** Study population characteristics with the EuReCa editions division. CPR, cardiopulmonary resuscitation; EMS, emergency medical services; EuReCa, European Registry of Cardiac Arrest; IQR, interquartile range

| **Characteristic** | **All editions**<br>**(n=8253)** | **EuReCa**<br>**One**<br>**(n=275)** | **EuReCa**<br>**Two (n=741)** | **EuReCa**<br>**Three**<br>**(n=7237)** | ***P -***<br>***value*** |
| --- | --- | --- | --- | --- | --- |
| Age (median;<br>IQR) | 68 (58 – 77) | 64(55 – 78) | 69(59 – 80) | 68(58 – 77) | 0.01 |
| Sex, male | 4758, 67.2% | 181, 66.1% | 467, 63.3% | 4110, 67.7% | 0.05 |
| CPR attempted by<br>EMS or bystander | n=8252<br>7440, 90.2% | n=274<br>133, 48.5% | n=741<br>382, 51.6% | n=7237<br>6925, 95.7% | 0.01 |
| CPR performed<br>with the support of<br>a dispatcher | n=1815<br>490, 27.0% | n=239<br>40, 16.7% | n=657<br>170, 25.9% | n=919<br>280, 30.5% | 0.01 |
| Shockable rhythm | n=7456<br>1560, 20.9% | n=238<br>26, 10.9% | n=729<br>67, 9.2% | n=6489<br>1467, 22.6% | 0.01 |

**Table II:** Clinical outcome in the EuReCa editions division. ROSC, any return of spontaneous circulation.

| **Characteristic** | **All editions** | **EuReCa**<br>**One** | **EuReCa**<br>**Two** | **EuReCa**<br>**Three** | ***P -***<br>***value*** |
| --- | --- | --- | --- | --- | --- |
| Any ROSC | n=7467<br>**1833, 24.5%** | n=131<br>**52, 39.7%** | n=728<br>**119, 16.3%** | n=6626<br>**1662, 25.2%** | 0.01 |
| 30 – days survival | n=7301<br>**547, 7.5 %** | n=48<br>**18, 37.5%** | n=98<br>**32, 32.7%** | n=7155<br>**497, 6.9%** | 0.01 |

Full CPR performed by bystander was associated with higher prevalence of ROSC as compared to the continuous chest compression only cardiopulmonary resuscitation (CCO CPR) and no CPR (26.0% vs. 23.3% vs. 21.7%; *P*=0.01). There was also an increase of ROSC incidence in the group where CPR was performed by bystander, although the precise technique of CPR performed was not comprehensively documented by EMS (26.2%; *P*=0.01). However, it is important to note that the ROSC rate is lower only in the case of patients where bystander CPR was not administered. Dispatcher-assisted cardiopulmonary resuscitation (TeleCPR or T-CPR) might be associated with increased ROSC incidence (22.1% vs 30.6%; *P* =0.01).

## DISCUSSION

In our study, we demonstrated that bystander-initiated CPR, particularly when supported a dispatcher, plays a pivotal role in improving immediate outcomes for OHCA in Poland. However based on the gathered data, there was a trend towards lower 30-day mortality without statistical significance both with and without dispatcher assistance.

### Survival factors

Several factors influence survival following OHCA, including patient characteristics, location of the event, and emergency response efficiency [3]. The cause of cardiac arrest was analyzed and coded based on the emergency medical team’s description, following the Utstein criteria. However, according to the literature, a substantial proportion of OHCA cases remain unexplained even after comprehensive diagnostic evaluations, with 12.3% of survivors having no identifiable underlying cause [11]. Klosiewicz et al. reported a ROSC rate of 54.9% [12], while Gräsner et al., in the EuReCa One study, highlighted variations depending on the country, ranging from 9.1% to 50%, and in the EuReCa Two study, reported an average ROSC rate of approximately 33% [13]. Among the possible variables in this category, cases of patients being transported to the hospital during ongoing CPR were also included. The EuReCa studies rely on emergency service reports, which may not fully capture all instances of ROSC. Some patients might not achieve ROSC at the scene/during transfer, but the patient could have achieved ROSC in the hospital, which partially explains the 30-day survival rate recorded in the National Death Registry.

### Dispatcher assisted CPR

In Poland, the activation time for ambulance services, defined as the time from receiving an emergency call to dispatching an ambulance, is a key factor in overall response time. Studies indicate that the average response time, including activation and travel, is 10.38 minutes [12]. The findings of our study align with previous research indicating that despite longer EMS response times in rural areas, the likelihood of bystander-initiated CPR is often higher than in urban settings. Cebula et al. demonstrated that while urban populations benefit from shorter EMS arrival times, rural communities compensate with higher rates of bystander intervention and dispatcher-assisted CPR [4]. These findings highlight the importance of integrating real-time guidance into prehospital care and AED mapping to strengthen the chain of survival in Poland. Performing CPR with the guidance and support of a medical dispatcher can greatly improve outcomes [14]. In Poland, both 112 (general emergency number) and 999 (ambulance service) [15] provide T-CPR instructions.

Currently, the 112-emergency call number operates across all EU Member States alongside national emergency numbers, ensuring universal access to emergency services. In Poland, 112 dispatchers handle all types of emergency calls (police, fire, and medical) and require general crisis management training, but may not have specialized medical qualifications. In contrast, 999 medical dispatchers must meet specific medical training standards, often including paramedic or emergency medical background and expertise, to provide accurate and structured TeleCPR instructions. Calls to 112 may be transferred to medical dispatchers, causing slight delays. However, in this study the type of emergency number answering the call was not collected. Our study suggests that T-CPR improves the likelihood of achieving ROSC (21.0% vs 27.4%; *P* =0.01). These findings highlight the critical impact of remote assistance in improving outcomes for cardiac arrest patients, particularly in situations where immediate, on-site medical intervention is limited. This likely results from the fact that guidance from a medical dispatcher during the resuscitation process helps to minimize uncertainties and facilitates the quicker execution of critical interventions. The study by Xu et al. demonstrated that T-CPR may reduce the time to first chest compression and other essential actions, such as opening the airway [16]. By providing instructions dispatchers improve the efficiency and effectiveness of the emergency response, leading to better patient outcomes during cardiac arrest situations. Certain studies suggested that the implementation of T-CPR might enhance survival rates, and in particular increasing prevalence of T-CPR appears to have a positive impact on the reduction of long-term neurological complications. Specifically, it has been observed to decrease the prevalence of brain complications from 40% to 32.1% [14]. Such findings underscore the potential of T-CPR to not only improve immediate survival outcomes, but also mitigate the risk of adverse neurological effects, contributing to a better quality of life for survivors. The evidence supports the critical role of T-CPR in improving outcomes in patients experiencing cardiac arrest.

### Locating caller in Poland

Timely initiation of high-quality CPR and early defibrillation are key factors influencing survival in OHCA [17]. Despite the increased availability and usage of AEDs in Poland, the absence of an updated AED map, as well as a longer delay before accurately locating a caller compared to most of Europe, decrease the dispatchers’ effectiveness. Advanced Mobile Location (AML) is being rolled out across most EU countries to provide emergency services with precise caller location data within 5 meters outdoors and 25 meters indoors. Poland is set to adopt AML by 2027, though its initial implementation faced significant public resistance, causing a delay. A new timeline for the rollout is now established [18].

After OHCA incidents, dispatchers, bystanders and device owners are not required to report AED usage [8]. The use of AEDs by either a bystander or a person sent to help is essential [19], especially in light of the systematic increase in frequency of shockable rhythm detection demonstrated in our study.

### Person sent to help

It should be noted that there are two different categories of “person sent to help”. One involving an institution registered in the “National Rescue and Firefighting System” [20] such as volunteer firefighters, Polish Red Cross rescue teams, mountain and water rescuers, and the other involving the growing community of the “First Rescuer” initiative. The presence of bag-valve masks or pocket masks increases the chance of performing full CPR compared to CCO CPR as data shows it might increase ROSC in the future. As is well known early CPR is crucial for survival, and our study showed an increase in CPR attempts, but evaluation of the potential impact of this increase is challenging, due to other factors depending on the entire survival chain. American studies of so-called HEARTSafe communities found no significant differences in 30-day survival rates between communities with and without first responder systems. Similarly, the EuReCa Three study in Poland, which evaluated bystander CPR, first responder actions, and AED use, found no significant correlation with 30-day survival. Although early CPR improves survival until hospital admission, it has not yet shown a difference in 30-day survival, highlighting the need for further research and system improvements. Our findings are consistent with those of Cone et al., who also found no significant differences in long-term survival outcomes despite increased survival until hospital admission with early CPR and T-CPR [21]

### Public Access to Defibrillation

The ideal treatment at the scene involves the use of on-site AED [22]. The concept of Public Access to Defibrillation (PAD), proposed by the American Heart Association (AHA) Task Force in 1995, aims to improve cardiac arrest survival rates. Data about AED usage was published as an observational study covering the period from 1 January 2008 to 31 December 2018, which analyzed 120 cases of AED use in public places in Poland, excluding emergency services [23]. The necessity of creating high-quality maps integrated with the dispatching system, and providing information regarding the availability of AEDs in the vicinity, is of paramount importance. Such improvements are likely to increase the use of AEDs by bystanders in cases of cardiac arrest in public places and on the street, although they may have a limited impact on their use in residences, where the incidence of OHCA is also significant. An analysis of existing AED location maps reveals that the majority of AEDs are situated in public buildings. Developing accurate and up-to-date maps showing the locations of AEDs in public spaces can significantly aid bystanders in quickly finding these life-saving devices. These maps should be accessible across various platforms, including mobile applications, websites, and via physical signage in public areas. Additionally, the integration of AED location data with navigation apps and emergency service platforms could prove crucial, as it can streamline the process of locating the nearest AED. Consideration should be given to planning and implementing solutions that could potentially increase the use of AEDs during cardiac arrests occurring in residence [22].

### Variability in ROSC outcomes

Our findings across the EuReCa studies show significant variability in ROSC outcomes. EuReCa One had the highest ROSC rates, while EuReCa Two saw a decline. In EuReCa Three, CPR attempts increased sharply but ROSC success was low, likely due to pandemic-related factors like bystander hesitation and EMS strain [24,25]. The incidence of any ROSC varied across the study periods, with 39.7% in EuReCa One, decreasing to 16.3% in EuReCa Two, and then rising to 25.2% in EuReCa Three (*P* = 0.01). In contrast, 30-day survival rates showed a marked and statistically significant decline, from 37.5% in EuReCa One and 32.7% in EuReCa Two to 6.9% in EuReCa Three (*P* = 0.01).The beneficial outcome in mortality in EuReCa Three might be related to a higher prevalence of CPR performed by bystander with or without dispatcher guidance as well as a higher rate of presence of shockable rhythm, which may be markers of more successful/faster resuscitation maneuvers. However, these findings may be also influenced by substantial differences in populations across the EuReCa One, Two, and Three studies, as well as variations in how the data was collected. This highlights the need for robust and standardized data collection methods like Utstein in future studies [26].

### Shockable rhythm

In Poland, EuReCa studies indicate a growing trend in the proportion of shockable rhythms among OHCA cases (Table I). Havranek et al. demonstrated that the presence of shockable rhythm significantly improves ROSC and survival outcomes in refractory OHCA, with 40% achieving favorable results compared to 5% with non-shockable rhythms [27]. These findings reflect progress in OHCA management and underscore the importance of early recognition and intervention such as early/high quality CPR etc. in improving survival rates [17,27].

### Impact of guidelines

Despite the substantial time gap between data collection periods in this study, the guideline updates introduced in 2015 and 2021 did not significantly impact the role of bystanders or dispatcher-assisted interventions in OHCA management. No major changes were implemented in the Polish system within routine clinical practice that would have substantially modified survival outcomes through bystander CPR or T-CPR, as reflected in the ERC recommendations during the studied period.

In summary, the growing involvement of bystanders in CPR has been pivotal, particularly with the support of tools like T-CPR, which are proven to enhance both survival rates and neurological outcomes for cardiac arrest patients. Systematic improvements in public AED availability, precise location mapping, and real-time dispatcher assistance also hold the potential to significantly boost survival rates in out-of-hospital cardiac arrest cases. Moreover, comprehensive evaluation of bystander and first responder interventions, coupled with continued research, is crucial to better understand the economic implications and long-term impacts of these efforts on survival and neurological health.

### Limitations

The most important limitation is the non-randomized design. Some unmeasured confounding variables might have an impact on the outcome. Experience and personal skills in CPR were also not incorporated in the analysis which might also be an important limitation. For instance, end-tidal COLJ (EtCOLJ) monitoring might serve as an indicator of the quality of chest compression and valuable prognostic tool during resuscitation for EMS, which might make it an interesting factor to analyze [28]. However routine use and systematic documentation in prehospital settings across Poland remain inconsistent.

Another major limitation is the lack of long-term follow-up of patients. For instance the influence of the duration and variant of mechanical ventilation or circulatory support was not included in the database.

Finally, the sample size might not be sufficient to detect the difference in mortality and adverse event rates. The observed differences in ROSC rates across EuReCa editions might partly result from differences in sample sizes.

It is important to note that ROSC refers to any restoration of circulation prior to hospital arrival, which may be transient and clinically limited. The short time frame of data collection limits the generalizability of the results, as it does not account for fluctuations in the incidence of OHCA over the course of a year, unlike in the POL_OHCA registry developed by Nadolny et al. [29]. However, this was a one-time initiative and included only predefined categories from official documents, rather than a comprehensive assessment based on the Utstein criteria. Another limitation is the 30-day survival rate calculation, which was based on the date of death from the national registry without accounting for the exact time of death. While we aimed to align ourselves with the Utstein criteria [26,30], this approach may introduce minor discrepancies in survival outcome classification. While EuReCa Three required digital and mandatory completion of emergency team reports, earlier studies (EuReCa ONE and TWO) relied on optional, paper based, and more accurate, documentation designed specifically for this research according to the Utstein criteria, which led to difficulties in data analysis [27].

One should also consider a potential bias caused by patients lost during follow-up. In Poland, the use of the Polish National Identification Number (PESEL) to monitor deaths excluded some patients who had not yet been assigned identification numbers, resulting in missing data and underrepresentation of cases in terms of 30-day survival. PESEL is a unique 11-digit national identification number assigned to all Polish citizens and residents. It is used for identity verification in administrative, legal, and medical systems. The number encodes the individual’s date of birth, gender, as well as an additional unique identifier. Moreover, paramedics often fill out records retrospectively, which introduces bias and affects the validity of key variables such as response times and performed interventions. Despite these limitations, our study reflects national practice based on a large, unselected patient cohort, utilizing the Utstein criteria to enable reliable comparisons with other healthcare systems. These criteria were applied through a detailed analysis of emergency medical team reports rather than relying solely on numerical data, ensuring a more comprehensive assessment of OHCA cases. This approach aligns with European standards for OHCA data collection and enables meaningful comparisons with other registries across the continent. Differences in emergency medical service structures, bystander intervention rates, and access to AEDs may influence survival outcomes, highlighting the need for further cross-national studies to evaluate the impact of healthcare system variations on OHCA management and prognosis.

## CONCLUSIONS

The results of this study stress the key role of bystanders and dispatchers in the chain of survival in OHCA cases. The results underscore the importance of improving the quality of CPR reporting in Poland to identify effective strategies and allocate resources efficiently. While the trends in the study are consistent with evidence from similar research in other countries, differences in prehospital systems and protocols may limit the direct applicability of these results. However, our study provides, for the first time in Poland, comprehensive insights into real-world clinical outcomes.

## Data Availability

All data produced in the present study are available upon reasonable request to the authors

## Notes

### Competing Interest Statement

The authors have declared no competing interest.

### Funding Statement

The study was funded from internal research funds allocated under the announcement of the Deputy Rector Representative for Science and International Cooperation of the Jagiellonian University Medical College for projects financed from the Ministry of Education and Science MEiN subsidy

### Summary of Updates

30 days survival was updated

## References

1. Maurer H, Masterson S, Tjelmeland IBM, Strömsöe A, Ortiz FR, Gräsner JT, et al. EuReCa – The European Registry of Cardiac Arrest and the related studies. Resusc Plus. 2024;19:100666.

2. Perkins GD, Graesner JT, Semeraro F, Olasveengen T, Soar J, Lott C, et al. European Resuscitation Council Guidelines 2021: Executive summary. Resuscitation. 2021;161:1–60.

3. Sip M, Puślecki M, Kłosiewicz T, Zalewski R, Dąbrowski M, Ligowski M, et al. A concept for the development of a pioneer regional out-of-hospital cardiac arrest program to improve patient outcomes. Kardiol Pol. 2020;78:875–81.

4. Cebula GM, Osadnik S, Wysocki M, Dyrda M, Chmura K, Nowakowski M, et al. Comparison of the early effects of out-of-hospital resuscitation in selected urban and rural areas in Poland. A preliminary report from the Polish Cardiac Arrest Registry by the Polish Resuscitation Council. Kardiol Pol. 2016;74:356–61.

5. Gach D, Nowak JU, Krzych ŁJ. Epidemiology of out-of-hospital cardiac arrest in the Bielsko-Biala district: A 12-month analysis. Kardiol Pol. 2016;74:1180–7.

6. Kłosiewicz T, Puslecki M, Zalewski R, Sip M, Perek B. Impact of automatic chest compression devices in out-of-hospital cardiac arrest. J Thorac Dis. 2020;12:2220–7.

7. Telec W, Baszko A, Dąbrowski M, Dąbrowska A, Sip M, Puslecki M, et al. Automated external defibrillator use in public places: A study of acquisition time. Kardiol Pol. 2018;76:181–5.

8. Żuratyński P, Ślęzak D, Dąbrowski S, Krzyżanowski K, Mędrzycka-Dąbrowska W, Rutkowski P. Use of public automated external defibrillators in out-of-hospital cardiac arrest in Poland. Medicina (Lithuania). 2021;57:298.

9. Orzechowski P, Kowalik I, Piotrowicz E. Feasibility of hybrid telerehabilitation as a component of the Managed Care after Acute Myocardial Infarction (MC-AMI) program in a 12-month follow-up: experience from a single center. Pol Arch Intern Med. 2023;29:133.

10. Gräsner JT, Wnent J, Lefering R, Herlitz J, Masterson S, Maurer H, et al. European registry of cardiac arrest study THREE (EuReCa THREE) – EMS response time influence on outcome in Europe. Resuscitation. 2025.

11. Mellor G. Unexplained cardiac arrest: Evolution within taxonomy? Polish Archives of Internal Medicine. Medycyna Praktyczna. 2019;129:148–50.

12. Kłosiewicz T, Skitek-Adamczak I, Zielinski M. Emergency medical system response time does not affect incidence of return of spontaneous circulation after prehospital resuscitation in one million central European agglomeration residents. Kardiol Pol. 2017;75:240–6.

13. Gräsner JT, Lefering R, Koster RW, Masterson S, Böttiger BW, Herlitz J, et al. EuReCa ONE—27 Nations, ONE Europe, ONE Registry: A prospective one month analysis of out-of-hospital cardiac arrest outcomes in 27 countries in Europe. Resuscitation. 2016;105:188–95.

14. Bagheri S, Sadeghi SM, Kazemi T, Nadimi E. Dispatcher-Assisted Bystander Cardiopulmonary Resuscitation (Telephone-CPR) and Outcomes after Out of Hospital Cardiac Arrest. Bull Emerg Trauma [Internet]. 2019 [cited 2025 Aug 9];7:307–13. www.beat-journal.com

15. Kogut B, Sitarz M. The Functioning of the e-Call System in Poland Compared to Other European Union Member States [Internet]. 2021 [cited 2025 Aug 9] European Research Studies Journal. 24;319–328. https://ersj.eu/journal/2720

16. Xu J, Qu M, Dong X, Chen Y, Yin H, Qu F, et al. Tele-Instruction Tool for Multiple Lay Responders Providing Cardiopulmonary Resuscitation in Telehealth Emergency Dispatch Services: Mixed Methods Study. J Med [Internet]. 2023 [cited 2025 Aug 9];25. https://www.jmir.org/2023/1/e46092

17. Kowalik R, Fojt A, Szczerba E, Peller M, Żukowska K, Opolski G. Impact of selected clinical factors on outcome of patients after out-of-hospital cardiac arrest treated with targeted temperature management. Pol Arch Intern Med [Internet]. 2019 [cited 2025 Aug 9];129:61–4. www.pamw.pl.

18. Announcement of the Marshal of the Sejm of the Republic of Poland dated April 10, 2024, regarding the publication of the consolidated text of the Act on National Medical Rescue Services. 2024.

19. Elhussain MO, Ahmed F k, Mustafa NM, Mohammed DO, Mahgoub IM, Alnaeim NA, et al. The Role of Automated External Defibrillator Use in the Out-of-Hospital Cardiac Arrest Survival Rate and Outcome: A Systematic Review. Cureus. 2023:15;e47721

20. Zuratynski P, Slezak D, Krzyzanowski K, Szczepanski R, Jaltuszewska S. State Medical Emergency System in Poland. Postępy Nauk Medycznych. 2019:32.

21. Cone DC, Burns K, Maciejewski K, Dziura J, McNally B, Vellano K. Sudden cardiac arrest survival in HEARTSafe communities. Resuscitation. 2020;146:13–8.

22. Folke F, Shahriari P, Hansen CM, Gregers MCT. Public access defibrillation: challenges and new solutions. Current Opinion in Critical Care. Lippincott Williams and Wilkins; 2023;28:168–74.

23. Ślęzak D, Robakowska M, Żuratyński P, Krzyżanowski K. Network of Automated External Defibrillators in Poland before the SARS-CoV-2 Pandemic: An In-Depth Analysis. Int J Environ Res Public Health. 2022;19.

24. Tokarek T, Dziewierz A, Malinowski KP, Rakowski T, Bartuś S, Dudek D, et al. Treatment Delay and Clinical Outcomes in Patients with ST-Segment Elevation Myocardial Infarction during the COVID-19 Pandemic. J Clin Med [Internet].2021 [cited 2025 May 15];10:3920. https://pmc.ncbi.nlm.nih.gov/articles/PMC8432080/

25. De Luca G, Dirksen MT, Spaulding C, Kelbæk H, Schalij M, Thuesen L, et al. Impact of diabetes on long-term outcome after primary angioplasty: insights from the DESERT cooperation. Diabetes Care [Internet]. 2012 [cited 2025 May 15];36:1020–5 https://europepmc.org/articles/PMC3609523

26. Grasner JT, Bray JE, Nolan JP, Iwami T, Ong MEH, Finn J, et al. Cardiac arrest and cardiopulmonary resuscitation outcome reports: 2024 update of the Utstein Out-of-Hospital Cardiac Arrest Registry template. Resuscitation. 2024;201.

27. Havranek S, Fingrova Z, Rob D, Smalcova J, Kavalkova P, Franek O, et al. Initial rhythm and survival in refractory out-of-hospital cardiac arrest. Post-hoc analysis of the Prague OHCA randomized trial. Resuscitation. 2022;181:289–96.

28. Paiva EF, Paxton JH, O’Neil BJ. The use of end-tidal carbon dioxide (ETCO2) measurement to guide management of cardiac arrest: A systematic review. Resuscitation. Elsevier Ireland Ltd; 2018;123:1–7.

29. Nadolny K, Zyko D, Obremska M, Wierzbik-Stroska M, Ladny JR, Podgorski M, et al. Analysis of out-of-hospital cardiac arrest in Poland in a 1-year Period: Data from the POL-OHCA registry. Kardiol Pol. 2020;78:404–11.

30. Idris AH, Bierens JJLM, Perkins GD, Wenzel V, Nadkarni V, Morley P, et al. 2015 revised Utstein-style recommended guidelines for uniform reporting of data from drowning-related resuscitation: An ILCOR advisory statement. Resuscitation. 2017;118:147–58.

